# Effects of exposure in virtual reality and in vivo on subjective symptom burden and physiological parameters of height-anxious healthy participants – A pilot study

**DOI:** 10.1101/2023.11.20.23298757

**Authors:** Vanessa Renner, Michael Witthöft, Jochen Hardt, Rupert Conrad, Katja Petrowski

## Abstract

Exposure in vivo is a highly effective treatment for anxiety disorders and acrophobia in particular. Nevertheless, it is rarely implemented in outpatient treatment. Exposure in virtual reality (VR) might be an alternative but its effect on subjective symptom burden and physiological parameters is questionable. Therefore, in this study, N = 33 participants with fear of heights received both in vivo and VR exposure in a randomized order. Subjective symptom burden before and after each exposure as well as heart rate and heart rate variability (SDNN, LF/HF ratio) during exposure sessions were assessed. Linear mixed models were calculated with the effect of condition (VR vs. in vivo) and scenario on HR, SDNN and LF/HF ratio. Subjective symptom burden was significantly reduced after both exposure sessions (p = .002; p < .001). Heart rate was significantly higher during exposure scenarios than baseline (p < .001). SDNN and LF/HF ratio reflected a significantly higher general power of HRV and a significantly higher activation of the sympathetic nervous system during exposure sessions compared to baseline and during VR exposure compared to in vivo. All in all, VR exposure shows comparable or superior effects compared to in vivo exposure regarding acrophobic fears and physiological parameters.

## Introduction

Acrophobia is characterized by an intense fear of heights, which is accompanied by body symptoms such as nausea, shivering, dizziness and trembling especially when confronted with heights. Individuals with acrophobia tend to avoid height-related situations such as high buildings or bridges (Coelho & Wallis, 2010). Cognitive behavioral therapy with exposure to anxiety-inducing situations in vivo (Lang et al., 2009) is considered as “gold standard” of psychotherapeutic treatment of anxiety disorders, as it shows consistently high therapeutic effects (Deacon & Abramowitz, 2004; Ost et al., 2004). Although exposure therapy is effective, it is rarely implemented in outpatient practice with only 13-17% of psychotherapists conducting exposure therapy (Pittig & Hoyer, 2017). Instead, CBT without in vivo exposure, interoceptive or in sensu exposure are mainly conducted (Hipol & Deacon, 2013; Klan & Hiller, 2014). A major obstacle for conducting in vivo exposure for treating acrophobia in particular are difficulties on the accessibility of high places, reducing the implementation of in vivo exposure. This could be solved by implementing exposure therapy in virtual reality (VRET). Past studies already assessed the effectiveness of VRET regarding subjective symptom burden. Therein, Eichenberg and Wolters (2012) showed that VRET is effective for social anxiety disorder, spider phobia, fear of flying, and fear of heights. When comparing VRET with in vivo exposure regarding agoraphobic symptom burden, comparable effects were shown for both treatment options (Emmelkamp et al., 2002; Krijn et al., 2004). Therefore, past studies suggest that VRET is an equally effective therapy procedure as in vivo exposure for acrophobia. Interestingly, these findings were found independent of the VR system used (Emmelkamp et al., 2002; Krijn et al., 2004). Since exclusively subjective anxiety was measured, the major lack of empirical evidence is the proof of effectiveness based on objective physiological data on anxiety symptoms.

Based on the emotional processing theory (EPT; Foa & Kozak, 1986), the patient’s fear network, consisting of propositions related to stimuli and phobic reactions, needs to be completely activated so exposure results in therapeutic change. Physiological activation can serve as an indicator for a successfully activated network (Foa & Kozak, 1986). A reliable and widely assessed measure of psychophysiological activation is the heart rate (HR) and the heart rate variability (HRV) as parameters of the autonomous nervous system. HR and HRV were not yet assessed in the past studies of VRET but were already assessed in studies investigating effects of in vivo exposure therapy in anxiety disorders (e. g. Alpers & Sell, 2008; Busscher et al., 2013). Therein it was found that HR measures at the beginning of the first exposure session predicted therapy outcome of claustrophobic patients whereas subjective symptom burden in the beginning did not predict therapy outcome (Alpers & Sell, 2008). Therefore, to receive a holistic picture of the effects of VRET and in vivo exposure in acrophobia, in this study subjective symptom burden and HR/HRV were assessed during an exposure session in VR and in vivo with height-anxious healthy participants.

## Method

### Sample

Sample size calculations were conducted using Stata 17.0 based on a power of 80 % and α = .05. Four points of measurement and two conditions (VR vs. in vivo) were assumed and matrices including expected effects per time point per condition were used for calculations. Based on the nature of the scenarios time effects were assumed with no differences between conditions (Carl et al., 2019; Wechsler et al., 2019). Based on this calculation, a sample size of N = 32 resulted. The recruitment was conducted via announcements at the Johannes Gutenberg-University Mainz, medical practices and online posts. Inclusion criteria were age between 18 and 65 years, ability to speak, read, write in German and giving informed consent and fear of height. Exclusion criteria were former or current mental disorder, cardiovascular diseases or other diseases influencing the cardiovascular system and intake of medication influencing the cardiovascular system. Interested participants were screened with the SCID-I screening of the DSM-IV (First & Gibbon, 2004; Wittchen et al., 2011). Therein, none of the participants reported to suffer from any past or current mental disorder. To assess the extent of fear of heights, interested participants were additionally screened with the Acrophobia Questionnaire (AQ; Cohen, 1977). This resulted in a final sample of N = 33 participants with fear of heights (60.6 % female). Age ranged from 19 to 35 years with M = 26.61 (SD = 4.18). 27.3% of the participants were students, 72.7% were unmarried. All of the participants provided written informed consent and the study procedure was conducted in accordance with the Declaration of Helsinki and was ethically approved by the Landesärztekammer Rheinland-Pfalz, Germany (2020-15411).

### Procedure

Participants fulfilling the described requirements were invited for one appointment lasting two to three hours. After giving informed consent, participants were introduced to the nature of fear, explanatory model of anxiety disorders and the idea of exposure. Before starting exposure sessions, participants answered the questionnaires and a three-minute resonance frequency breathing was implemented with a 5 x 5 interval (5 seconds inhale, 5 seconds exhale) as baseline HRV measure (scenario 1). In this within-design, participants received both exposure procedures (VR and in vivo) in a randomized order. After the first exposure session, participants filled in questionnaires and then started the second exposure followed by answering questionnaires again.

The VR exposure session was the freely available programmed scenario Richie’s Plank Experience (https://www.oculus.com/experiences/quest/1642239225880682/?locale=de_DE). Therein, participants can use an elevator in a skyscraper and can choose to stop at a plank. When this button is chosen, the elevator goes up and when the door opens, participants see a wooden plank on a top of a skyscraper on which they can walk. In our study, at first, participants spend time at the city where they can walk in small, limited space (scenario 2). Next, they step into the elevator and choose “plank” and the elevator goes up. Participants were asked to walk on this plank and stay at the very end of it (scenario 3). Lastly, participants were asked to leave the plank, going down with the elevator back to the city where the scenario started (scenario 4). In vivo, the procedure was parallelized by also implementing three scenes. First, participants went out on the roof of a five-storey building where they went on the edge and looked down to the ground (scenario 2). Next, they walked in the middle of the roof with a clear view of the surrounding (scenario 3). Last, the walked back to the door and left the roof (scenario 4).

### Clinical Assessment

The agoraphobic cognitions questionnaire (ACQ; Chambless et al., 1984; Ehlers et al., 2001) was handed out to the participants before and after each exposure measuring fearful panic beliefs and catastrophic cognitions about panic as a self-report. 15 items need to be answered on a 5-point scale ranging from 0 (thought never occurs) to 4 (thought always occurs). The Acrophobia Questionnaire (AQ; Cohen, 1977) consists of 20 items assessing fear of height-related situations such as bridges, balconies or ferries wheels. Participants rate their fear on a scale from 0 (not anxious at all) to 6 (very anxious).

After VR exposure sessions, participants answered the Igroup Presence Questionnaire (IPQ; Schubert, 2003) and the Simulation Sickness Questionnaire (SSQ; Neukum & Grattenthaler, 2006). The IPQ (Schubert, 2003) measures the sense of presence experienced in a virtual environment. It includes three subscales (spatial presence, involvement, experienced realism), which can be combined into an overall score. The SSQ (Neukum & Grattenthaler, 2006) assesses sickness symptoms such as headache, nausea or dizziness elicited by VR. Participants rate the occurrence of 16 different symptoms on a scale from 0 (no perception) to 3 (severe perception). Scores for nausea, oculomotor disturbance, disorientation and total simulator sickness can be computed.

### Technical devices and heart rate variability

For VR exposure, a standalone Oculus Quest by Meta was used with the freely available program ‘Richie’s Plank Experience’ (https://www.oculus.com/experiences/quest/ 1642239225880682/?locale=de_DE). Participants interacted with their hands with the environment (e. g. pressing the button in the elevator) and did not use remote controller. HRV was obtained with a Polar Watch V800 assessing HR and HRV with a chest strap. To eliminate extra beats or erroneous values of the R–R interval data, the software Polar ProTrainer 5 (Polar, Germany) was used to post-process the recordings by an automatic filtering process method (filter power: moderate, minimum protection zone: 6 sqm). The time-based and frequency-based HRV parameters standard deviation of all NN intervals (SDNN) and the ratio of power in low-frequency range/power in high-frequency range (LF/HF ratio) were calculated. SDNN was used to assess the overall power of the HRV (ESC and NASPE 1996). The parameter LF/HF indicates the interaction between the sympathetic and parasympathetic nervous system of the autonomous nervous system. Analyses of the parameters were conducted with Kubios HRV Standard (Tarvainen et al., 2014). HR and HRV parameters were transformed using logarithm naturalis.

### Statistical analyses

Repeated measures ANOVA were calculated to assess differences regarding subjective symptom burden (ACQ, AQ). Greenhouse-Geisser correction was used when necessary. For comparison of HR and HRV parameters, VR and in vivo exposure were split into four scenarios each: First scenario is the resonance frequency breathing before both exposures. In VR, it is followed by being in the city and going up with the elevator (scenario 2), staying at the plank (scenario 3) and going back down with the elevator to the city (scenario 4). During in vivo exposure, standing on the roof, walking on the edge and looking down to the ground is scenario 2. Followed by walking in the middle of the roof with a clear view of the surrounding (scenario 3) and walking back to the door and leaving the roof (scenario 4). Additionally, each scenario was split into two slots: first 30 seconds in the beginning and last 30 seconds in the end of each scenario. Except for resonance frequency breathing, peak HR and HRV parameters were also calculated. Linear mixed models (LMM) as recommended when analyzing nested data (Hox et al., 2017) were calculated using Jamovi 2.3 (The Jamovi Project, 2020). The effect of condition (VR vs. in vivo), scenarios and slot (first 30 seconds, peak, last 30 seconds) on HR, SDNN and LF/HF ratio serving as dependent variables was assessed. A random effect for participants (random-intercept term) to allow the intercept to vary across participants was included. Three models were calculated, one for each HRV parameter. Non-significant interactions within the models were excluded stepwise, highest p-values were excluded first.

## Results

There was a significant effect for time regarding the ACQ when comparing scores before and after exposure sessions (VR: M_pre_ = 1.68, SD_pre_ = 0.46; M_post_ = 1.64, SD_post_ = 0.51; in vivo: M_pre_ = 1.76, SD_pre_ = 0.46; M_post_ = 1.49, SD_post_ = 0.32; F(1, 32) = 11.62, p = .002, η_p_^2^ = .266) but no significant effect for condition (VR vs. in vivo; F(1, 32) = 1.20, p = .281). A significant interaction for time x condition was found (F(1, 32) = 12.27, p = .001, η_p_^2^ = .277). The AQ showed significant pre-/post-differences (VR: M_pre_ = 1.43, SD_pre_ = 0.57; M_post_ = 1.20, SD_post_ = 0.59; in vivo: M_pre_ = 1.49, SD_pre_ = 0.55; M_post_ = 1.31, SD_post_ = 0.59; F(1, 32) = 13.84, p < .001, η_p_^2^ = .302), reflecting a symptom reduction after exposure sessions compared with before exposure sessions. A significant effect of condition (F(1, 32) = 4.34, p = .045, η_p_^2^ = .119) was found with higher pre/post values for in vivo exposure.

After VR exposure, the mean IPQ score was M = 2.62 (SD = 0.36) ranging from M = 2.62 to M = 4.00. A total of 57.6% did report a general sense of presence in the VR environment of at least M = 3.46. The mean score of spatial presence was M = 3.30 (SD = 0.59), of involvement M = 3.17 (SD = 0.42) and experienced realism M = 3.61 (SD = 0.87). Regarding the SSQ, the most problems occurred regarding nausea (M = 3.48, SD = 2.46) followed by oculomotor disturbances (M = 3.30, SD = 3.07) and disorientation (M = 2.64, SD = 2.78). In total, 6.1% did not report any symptoms of simulation sickness, 72.7% of the participants scored smaller than 10. The maximum score on the SSQ was 20 out of 48.

Regarding HR, there was a significant main effect of scenario (F(3, 672.00) = 22.97, p < .001) and slot (F(2,672.00) = 35.32, p < .001), but not for condition (F(1, 672.00) = 0.46, p = .500), indicating that VR and in vivo exposure showed comparable effects on HR. Additionally, significant interactions were found for condition x scenario (F(3,672.00) = 6.16, p < .001), scenario x slot (F(5, 672.00) = 9.90, p < .001) and condition x scenario x slot (F(5, 672.00) = 3.04, p = .010). Fixed effects parameter estimates for HR can be derived from table 1.

**Table 1.**
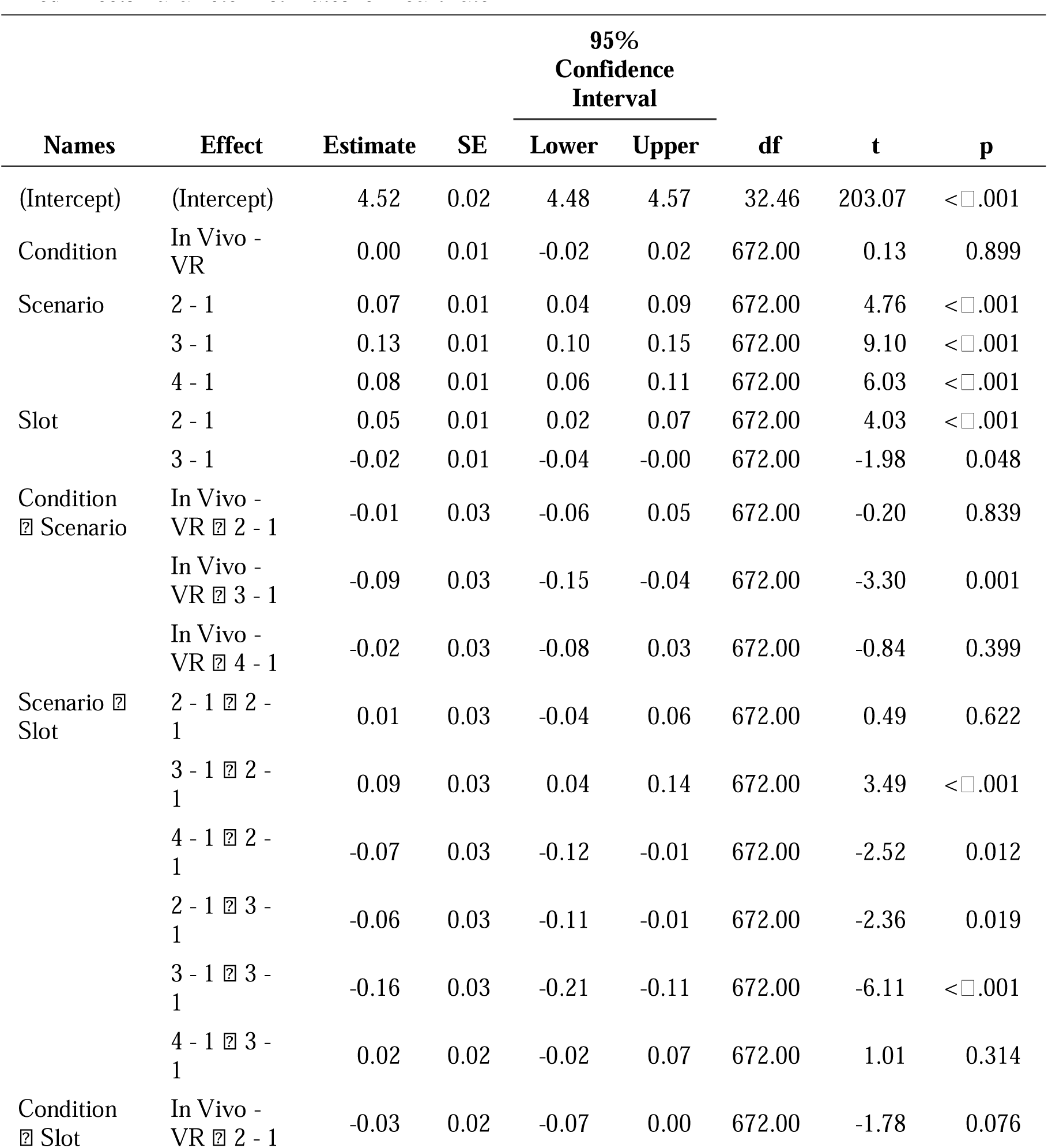
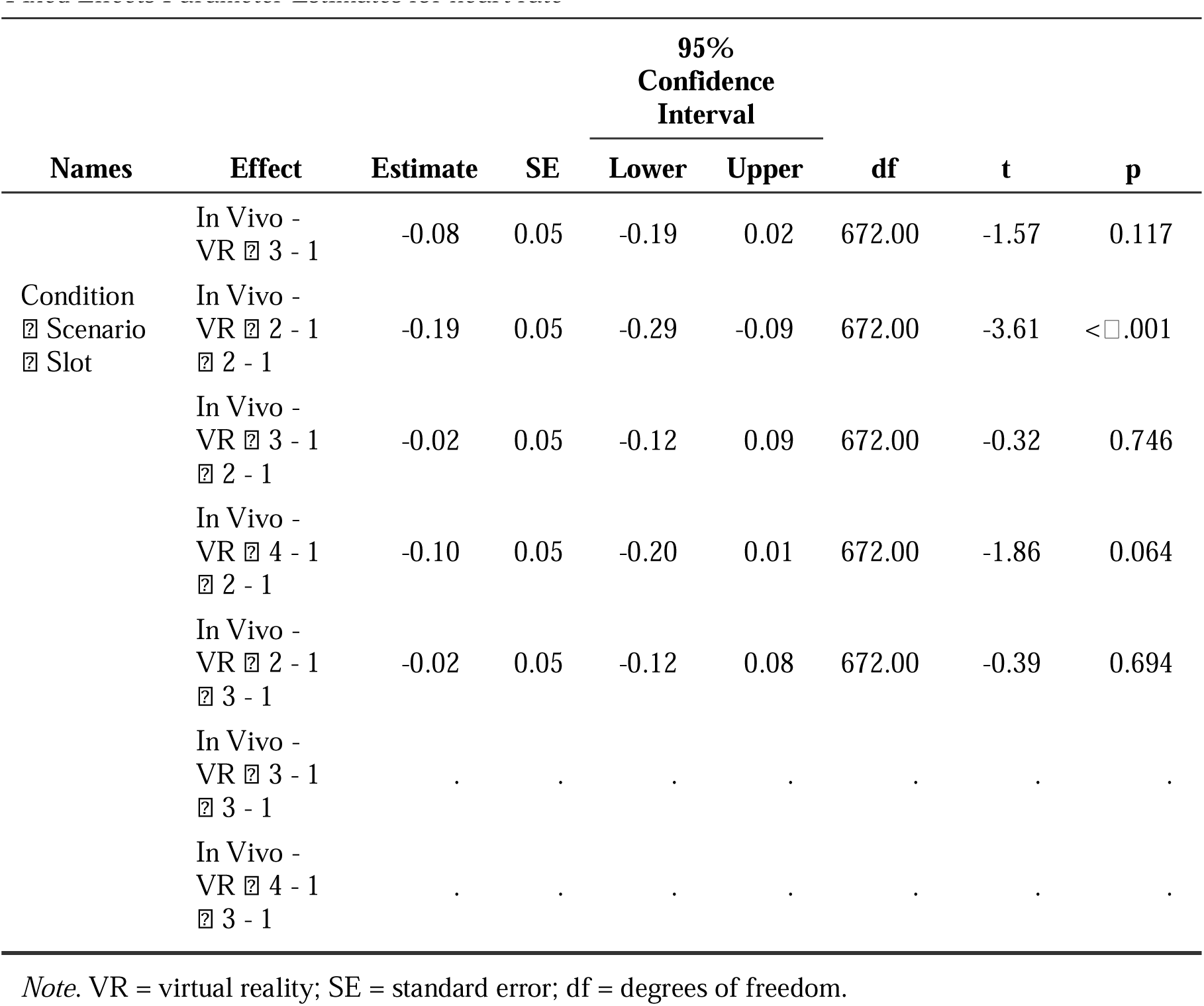
Fixed Effects Parameter Estimates for heart rate

Regarding SDNN, significant main effects were found for condition (F(1, 679.00) = 4.76, p = .030) and scenario (F(3,679.00) = 18.36, p < .001) but not for slot (F(2, 679.00) = 0.46, p = .631), reflecting higher SDNN in VR than in vivo exposure and differential effects of the presented scenarios on SDNN. Significant interactions were found for condition x scenario (F(3, 679.00) = 3.28, p = .021) and scenario x slot (F(5, 679.00) = 3.82, p = .002). Fixed effects parameter for SDNN can be derived from table 2.

**Table 2.**
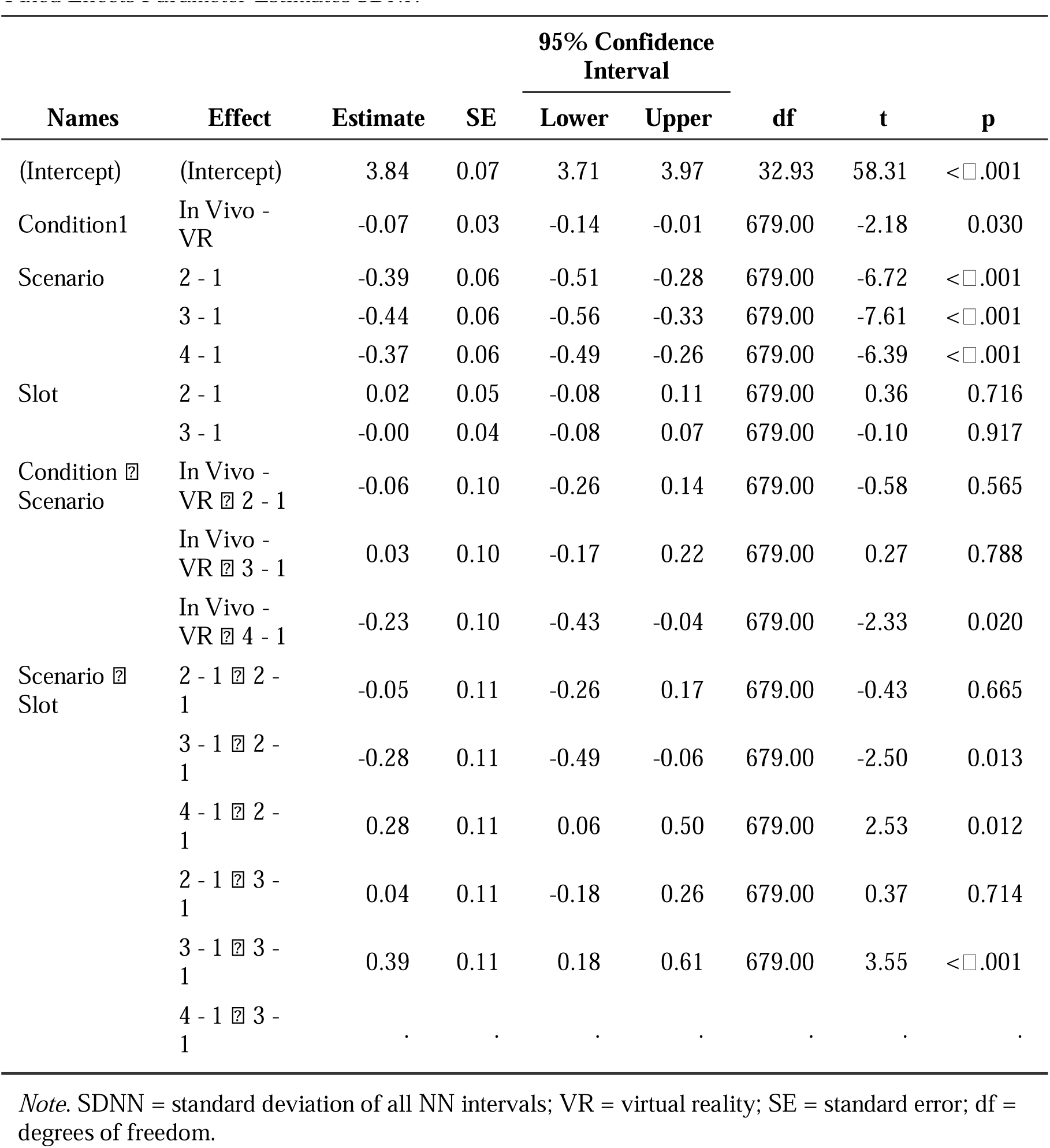
*Fixed Effects Parameter Estimates SDNN*

| Names | Effect | Estimate | SE | 95% Confidence Interval |  | df | t | p |
| --- | --- | --- | --- | --- | --- | --- | --- | --- |
|  |  |  |  | Lower | Upper |  |  |  |
| (Intercept) | (Intercept) | 3.84 | 0.07 | 3.71 | 3.97 | 32.93 | 58.31 | < .001 |
| Condition1 | In Vivo - VR | -0.07 | 0.03 | -0.14 | -0.01 | 679.00 | -2.18 | 0.030 |
| Scenario | 2 - 1 | -0.39 | 0.06 | -0.51 | -0.28 | 679.00 | -6.72 | < .001 |
|  | 3 - 1 | -0.44 | 0.06 | -0.56 | -0.33 | 679.00 | -7.61 | < .001 |
|  | 4 - 1 | -0.37 | 0.06 | -0.49 | -0.26 | 679.00 | -6.39 | < .001 |
| Slot | 2 - 1 | 0.02 | 0.05 | -0.08 | 0.11 | 679.00 | 0.36 | 0.716 |
|  | 3 - 1 | -0.00 | 0.04 | -0.08 | 0.07 | 679.00 | -0.10 | 0.917 |
| Condition × Scenario | In Vivo - VR × 2 - 1 | -0.06 | 0.10 | -0.26 | 0.14 | 679.00 | -0.58 | 0.565 |
| | In Vivo - VR × 3 - 1 | 0.03 | 0.10 | -0.17 | 0.22 | 679.00 | 0.27 | 0.788 |
| | In Vivo - VR × 4 - 1 | -0.23 | 0.10 | -0.43 | -0.04 | 679.00 | -2.33 | 0.020 |
| Scenario × Slot | 2 - 1 × 2 - 1 | -0.05 | 0.11 | -0.26 | 0.17 | 679.00 | -0.43 | 0.665 |
| | 3 - 1 × 2 - 1 | -0.28 | 0.11 | -0.49 | -0.06 | 679.00 | -2.50 | 0.013 |
| | 4 - 1 × 2 - 1 | 0.28 | 0.11 | 0.06 | 0.50 | 679.00 | 2.53 | 0.012 |
| | 2 - 1 × 3 - 1 | 0.04 | 0.11 | -0.18 | 0.26 | 679.00 | 0.37 | 0.714 |
| | 3 - 1 × 3 - 1 | 0.39 | 0.11 | 0.18 | 0.61 | 679.00 | 3.55 | < .001 |
| | 4 - 1 × 3 - 1 | . | . | . | . | . | . | . |

Note. SDNN = standard deviation of all NN intervals; VR = virtual reality; SE = standard error; df = degrees of freedom.

For LF/HF ratio, significant main effects for condition (F(1, 689.00) = 5.18, p = .023) and scenario (F(3, 689.00) = 17.19, p < .001) were found but no significant interactions. Fixed effects parameter estimates for LF/HF ratio can be derived from table 3.

**Table 3.**
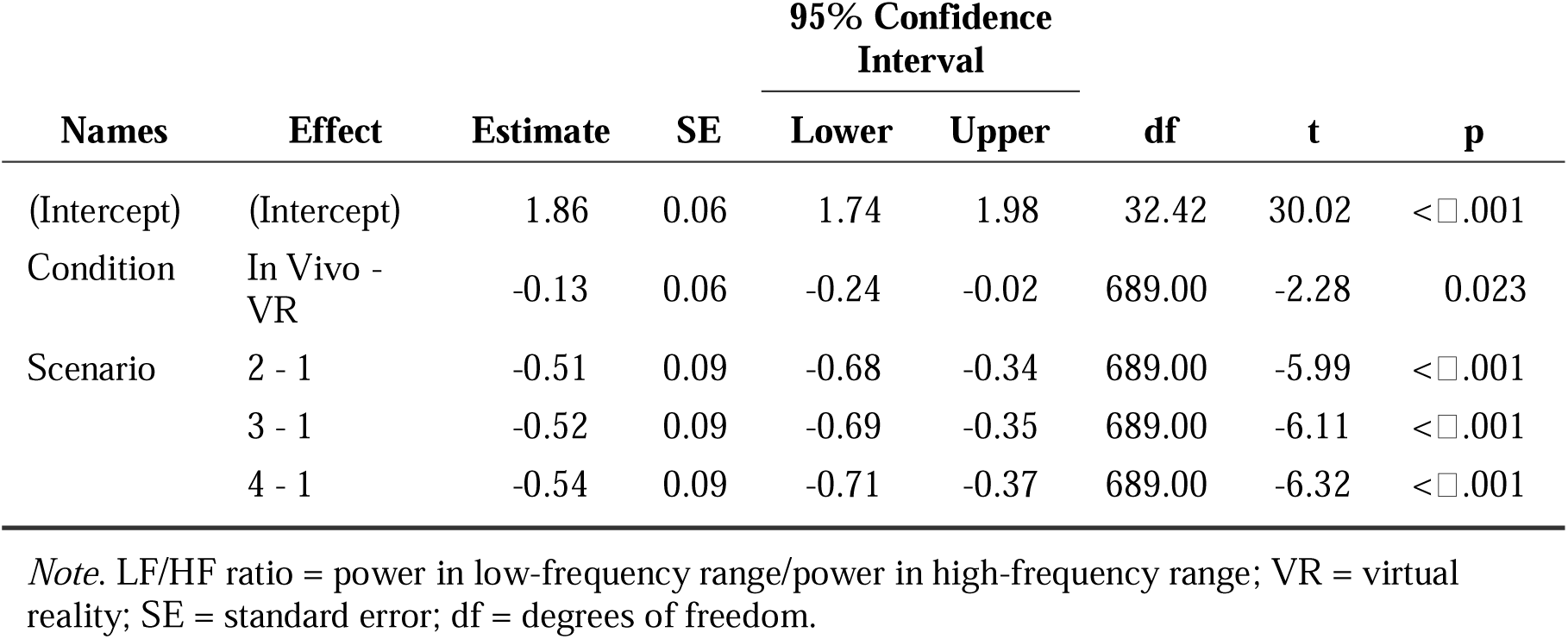
Fixed Effects Parameter Estimates for LF/HF ratio

| Names | Effect | Estimate | SE | 95% Confidence Interval |  | df | t | p |
| --- | --- | --- | --- | --- | --- | --- | --- | --- |
|  |  |  |  | Lower | Upper |  |  |  |
| (Intercept) | (Intercept) | 1.86 | 0.06 | 1.74 | 1.98 | 32.42 | 30.02 | <□.001 |
| Condition | In Vivo - VR | -0.13 | 0.06 | -0.24 | -0.02 | 689.00 | -2.28 | 0.023 |
| Scenario | 2 - 1 | -0.51 | 0.09 | -0.68 | -0.34 | 689.00 | -5.99 | <□.001 |
|  | 3 - 1 | -0.52 | 0.09 | -0.69 | -0.35 | 689.00 | -6.11 | <□.001 |
|  | 4 - 1 | -0.54 | 0.09 | -0.71 | -0.37 | 689.00 | -6.32 | <□.001 |

*Note.* LF/HF ratio = power in low-frequency range/power in high-frequency range; VR = virtual reality; SE = standard error; df = degrees of freedom.

## Discussion

This is the first study that investigated highly standardized and controlled the effect of VR versus in vivo exposure on a subjective as well as psychophysiological level. The effects of VR and in vivo exposure on subjective symptom burden and HR/HRV were assessed in height-anxious healthy participants. Regarding subjective symptom burden, agoraphobic cognitions and subjective fear of heights was significantly reduced by VR as well as by in vivo exposure. Regarding physiological measures, exposure scenarios led to a stronger activation reflected by higher HR compared to baseline. HR levels showed a highly activated autonomous nervous system during the main exposure scenario in VR when participants stood on a plank.

SDNN and LF/HF ratio were significantly higher during VR exposure than during in vivo exposure reflecting a higher overall power of HRV and a stronger activation of the sympathetic nervous system. In VR scenario 4, the SDNN reached a peak compared to the other scenarios. This is probably due to participants leaving the plank in the beginning of this section leading to a high activation of the autonomous nervous system. HR was also significantly reduced at the end of each exposure scenario compared to the beginning (slot 1 vs. 3). This could be due to a habituation process within the scenarios wherein physiological activation decreased. All in all, this study shows, in line with results of past studies (Carl et al., 2019; Wechsler et al., 2019), comparable or even superior effects of VR and/to in vivo exposure on acrophobic fear and physiological measures of HR and HRV in participants with fear of heights.

The current findings are in line with past studies showing comparable effects of VRET and in vivo exposure therapy on subjective symptom burden (Emmelkamp et al., 2002; Krijn et al., 2004) wherein subjective symptom burden was reduced after VRET. Beyond previous studies, this study also shows effects of VR on physiological parameters. Also, regarding objective measures, VR shows comparable or even superior effects compared to in vivo exposure. Therefore, VR exposure sessions seem to be an at least equally effective option as in vivo exposure. Despite the high effectiveness of in vivo exposure, as described above, it is rarely implemented in outpatient treatment (Pittig & Hoyer, 2017). Especially in outpatient treatment where in vivo exposure is rarely implemented, VR exposure could be an easily applicable exposure treatment. This is of special interest when implementation of in vivo exposure is not possible or associated with high effort and constraints as in acrophobia or agoraphobia. To evaluate effects of an exposure therapy for acrophobic or agoraphobic patients, future studies should implement an exposure therapy in VR and in vivo with several exposure sessions comparing subjective symptom burden and physiological measures. This would give further insights in the effectiveness of VR exposure and its potential for an addition to or lower-threshold alternative for exposure therapy in vivo. It would also be of interest to further assess interactions between subjective symptom burden and physiological measures such as HR/HRV and its predictive role for therapy outcome.

Despite its strengths, this study also shows some limitations. Even though the sample did show symptoms of acrophobia this was a sample with subjectively lower levels of fear of height assessed via the AQ. Therefore, the effectiveness of VRET has to be shown in a patient sample with more severe or chronic symptoms. Nevertheless, even in this sample, both exposure sessions did reduce fear of height significantly. In the present study only one session of VR and in vivo exposure was conducted. Also, long-term effects were not investigated in this study but should be assessed in the future.

The present study shows similar to slightly superior effects of VR and in vivo exposure on subjective symptom burden and autonomous nervous system activity in healthy participants with fear of heights. This emphasizes the relevance of taking VR exposure into account for an addition to or alternative for in vivo exposure, which is rarely implemented in outpatient treatment. An easy-to-implement and equally effective treatment could increase the implementation of exposure and therefore optimize the treatment quality for acrophobic patients in particular and patients with anxiety disorders in general.

## Data Availability

All data produced in the present study are available upon reasonable request to the authors

## Acknowledgements

We would like to thank the medical doctorates Lukas Schäfer and Alan Karimi for their dedicated efforts and great support in data collection and processing as well as all of the participants for taking part in this study.

The authors received no funding from an external source.

---

**Figure 1.**
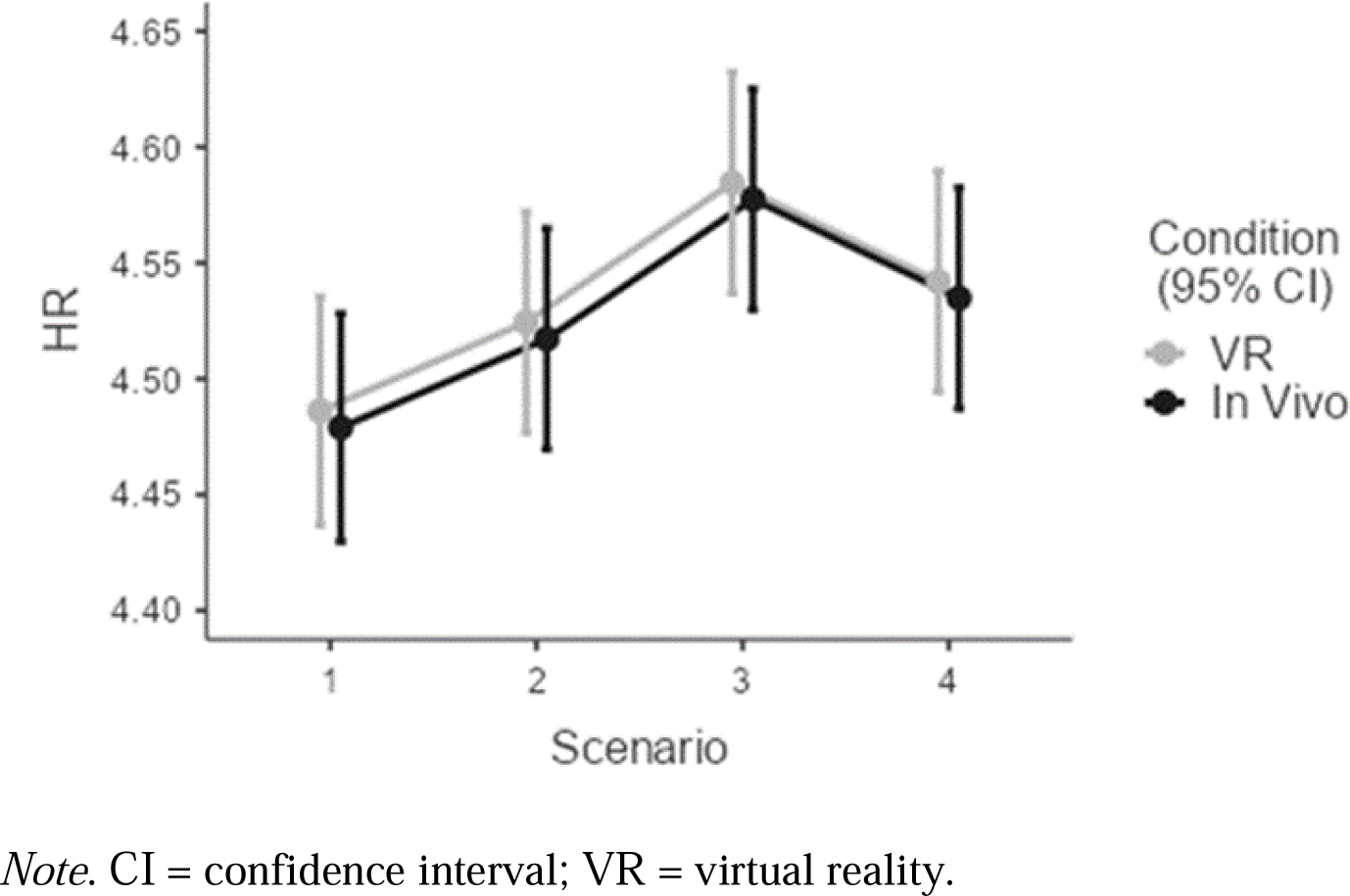
Heart rate for each scenario during VR and in vivo exposure

**Figure 2.**
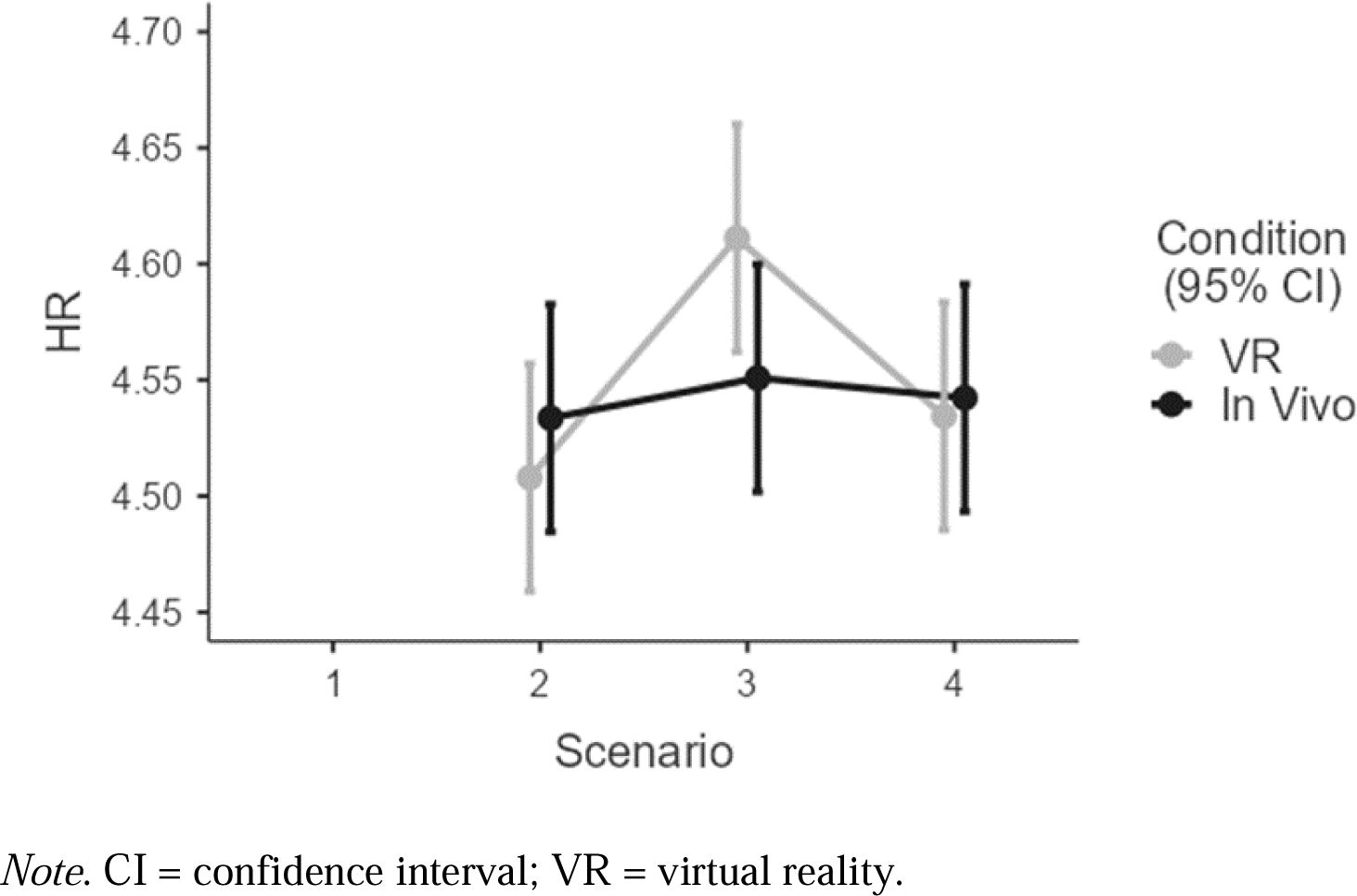
Interaction effects of heart rate

**Figure 3.**
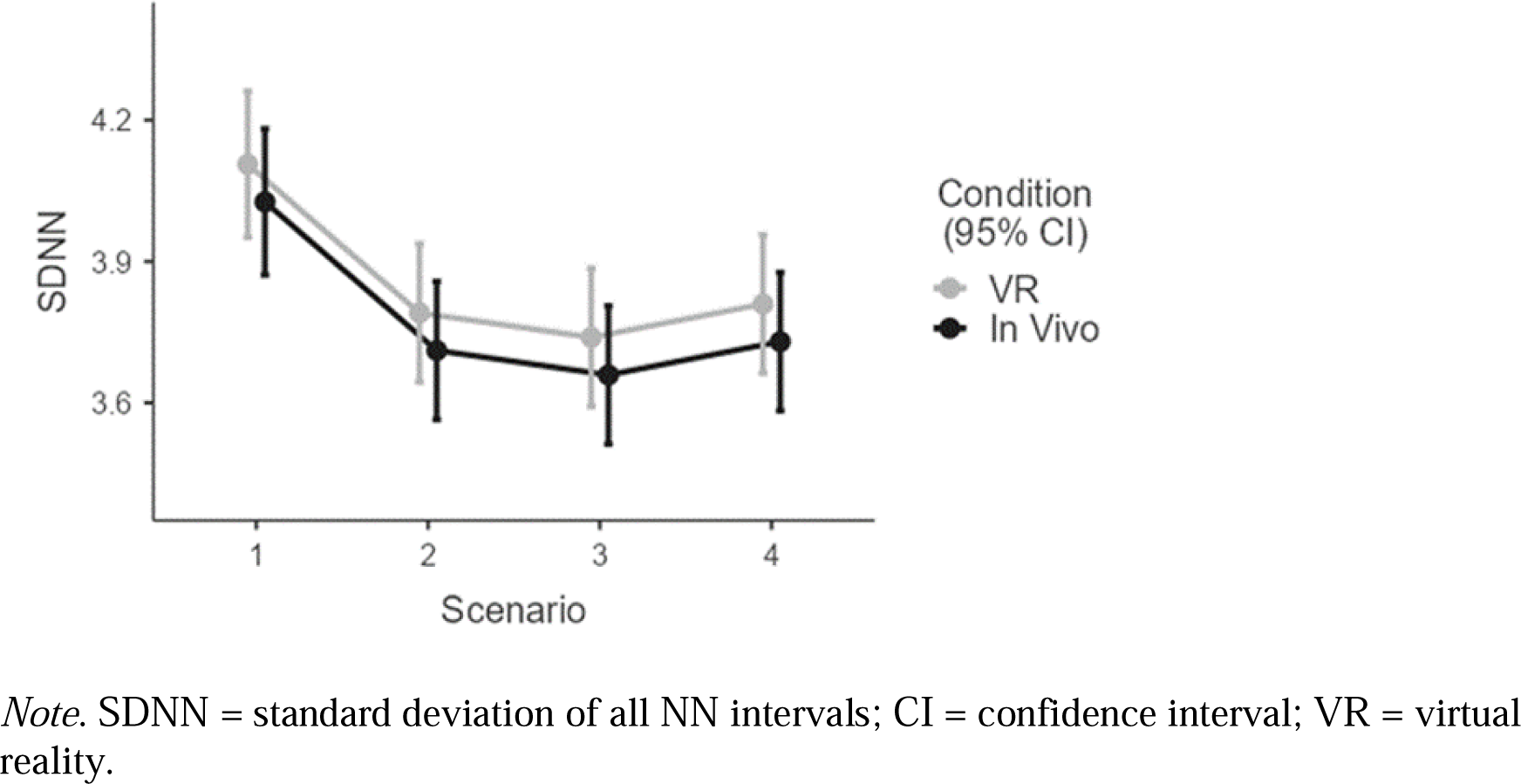
SDNN for each scenario during VR and in vivo exposure

**Figure 4.**
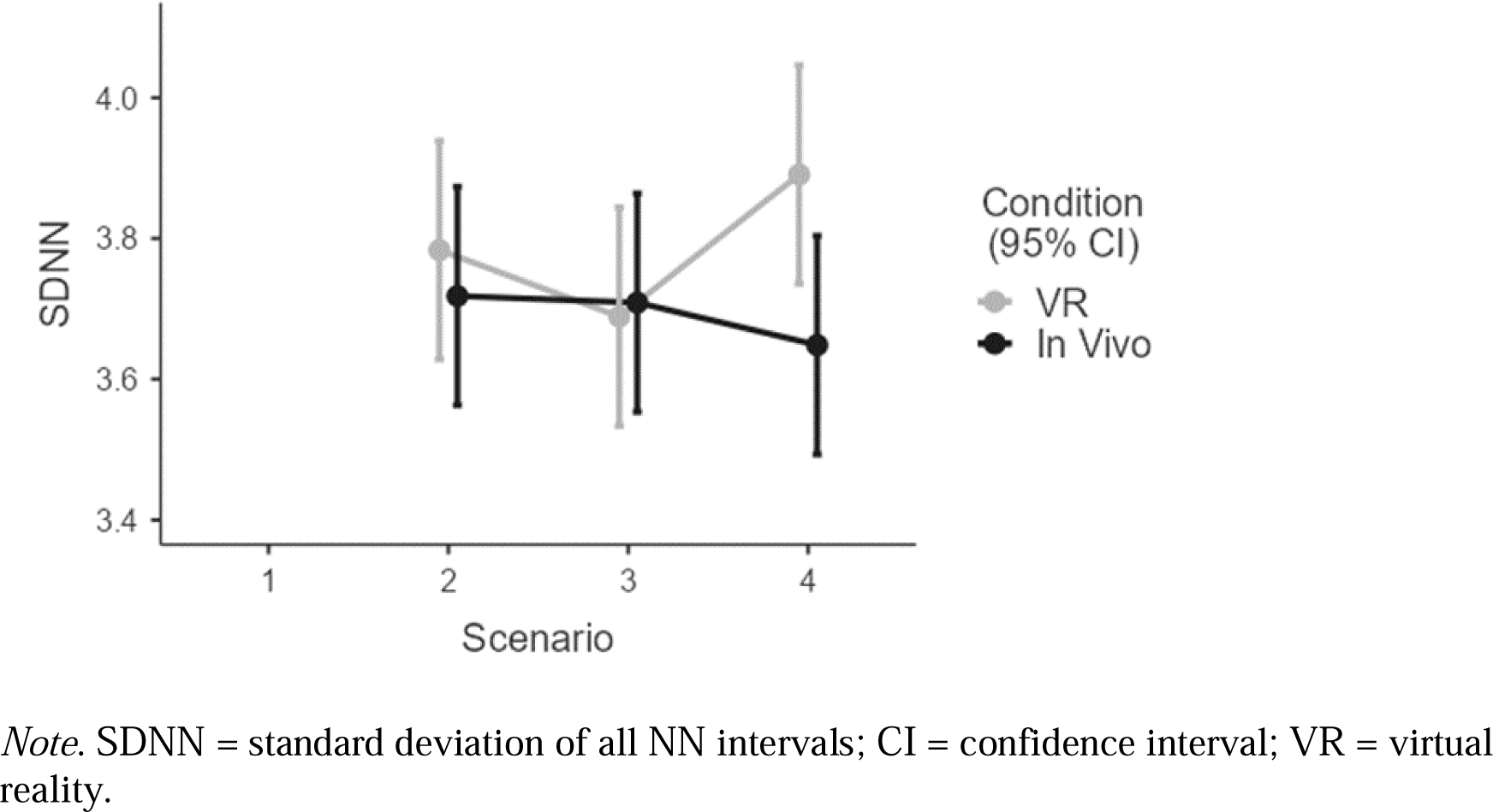
Interaction effects of SDNN

**Figure 5.**
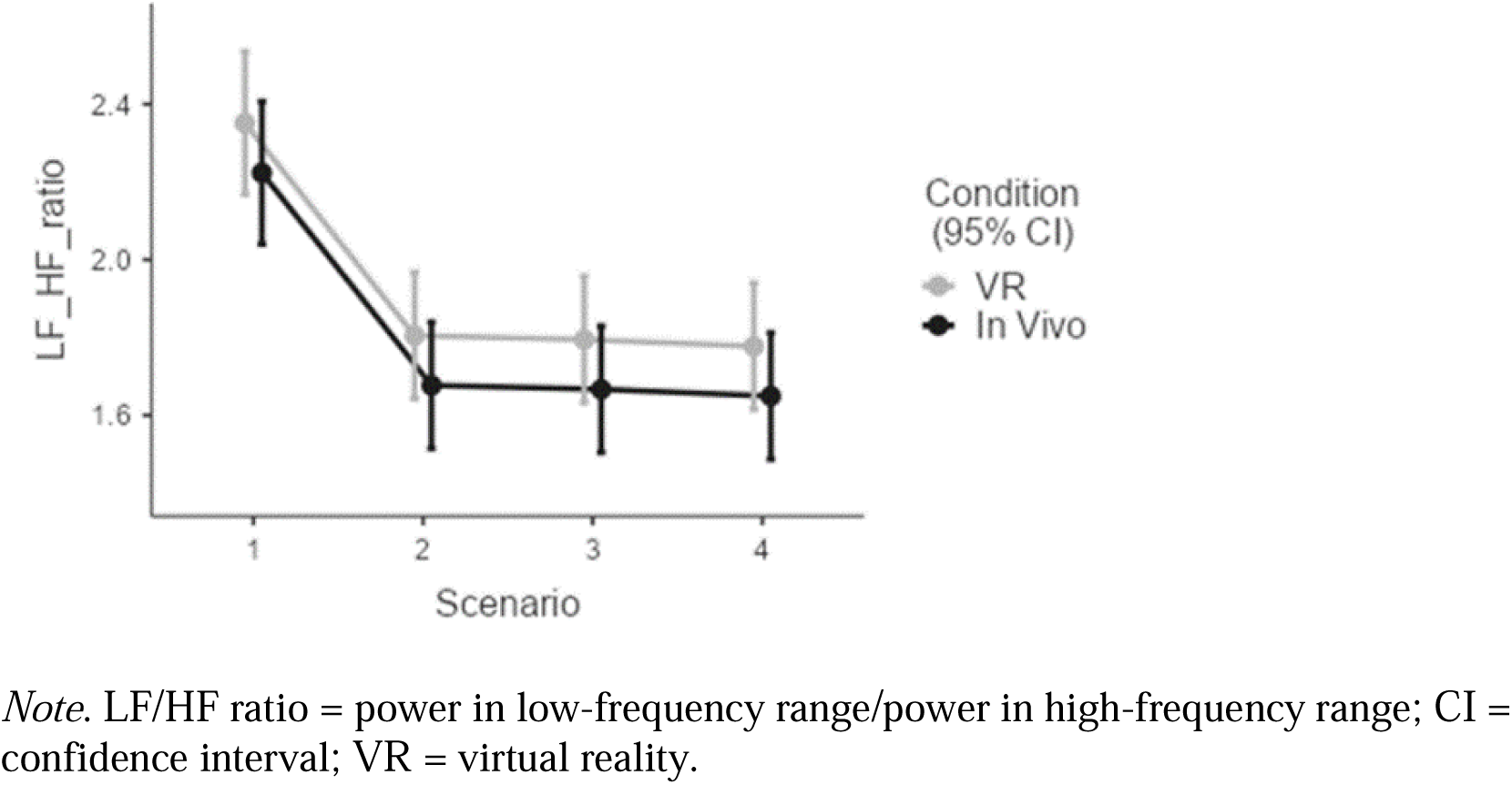
LF/HF ratio for each scenario during VR and in vivo exposure

